# High-level anxiety is associated with worse clinical symptoms and aberrant brain networks in Parkinson’s disease

**DOI:** 10.1101/2023.09.25.23296076

**Authors:** Zhichun Chen, Guanglu Li, Liche Zhou, Lina Zhang, Jun Liu

**Author notes:** **Correspondence:** Jun Liu, Department of Neurology and Institute of Neurology, Ruijin Hospital affiliated to Shanghai Jiao Tong University School of Medicine, Shanghai, 200025, China. E-mail address (J. Liu).

## Abstract

**Background:** Anxiety is one of the most common psychiatric symptoms in Parkinson’s disease (PD). Whether anxiety shapes disease progression and brain network aberrations in PD remain largely unknown. The objective of present study is to investigate whether anxiety exacerbates clinical symptoms and brain network abnormalities of PD patients.

**Methods:** PD patients performing magnetic resonance imaging from Parkinson’s Progression Markers Initiative (PPMI) database were specifically included. According to the scores of State-Trait Anxiety Inventory (STAI), PD patients were classified into lower quartile group (STAI score rank: 0%∼25%), interquartile group (STAI score rank: 26%∼75%), and upper quartile group (STAI score rank: 76%∼100%) based on their STAI score quartiles to investigate how anxiety affects clinical manifestations and brain networks.

**Results:** Anxiety was independently associated with both motor and non-motor symptoms of PD patients. Consistently, PD patients in the upper quartile group showed more severe non-motor symptoms compared to lower quartile group. Moreover, they also exhibited significantly different topological metrics in structural network compared to lower quartile group. Furthermore, we demonstrated that differential network metrics mediated the associations between anxiety and motor and non-motor symptoms of PD patients.

**Conclusions:** PD patients with higher anxiety level exhibited more severe clinical manifestations and disruptions of brain network topology. Diverse structural network metrics were associated with motor and non-motor symptoms of PD patients.

## Introduction

Parkinson’s disease (PD) is a chronic and progressive neurodegenerative disorder affecting millions of populations worldwide ^1,2^. It is characterized by classical motor symptoms such as bradykinesia, resting tremor, rigidity, postural instability and concomitant non-motor symptoms including neuropsychiatric complications like anxiety, depression, and insomnia ^1–3^. Anxiety is a severe non-motor symptom for more than one-third of PD patients and has extremely negative impact on quality of life ^3–8^. Anxiety occurs mostly in the “off” state and its frequency tends to fluctuate ranging from 3.1% to 67.7% in PD patients with motor fluctuations ^9^. According to previous studies, anxiety was associated with multiple risk factors, including older age, female, motor subtypes, motor fluctuations, disease duration, autonomic dysfunction, worse cognitive functioning, depression, REM sleep behavior disorder (RBD), poor sleep quality, insomnia, and olfactory loss ^10–15^. Anxiety significantly exacerbates clinical symptoms and contributes to worse disease outcomes ^16–21^. Specifically, anxiety is associated with worse motor manifestations and complications, including freezing of gait ^18,19,22,23^, impaired motor control ^17^, dyskinesias and motor fluctuations ^16^. Anxiety is also significantly associated with more severe non-motor symptoms, such as cognitive impairment ^20,21,24^, sleep disorders ^8,10^, physical pain ^25^, frailty ^25^, impulse control disorders ^26,27^, autonomic dysfunction ^11,28^, and depression ^5,28,29^. Therefore, anxiety is an important non-motor symptom significantly shaping disease severity and outcomes ^7,30^.

Because anxiety plays a crucial role in PD, it is imperative to thoroughly examine and understand the molecular mechanism and pathophysiological pathways linked to the occurrence of anxiety in PD ^7,30,31^. However, currently, the exact neural underpinnings for anxiety in PD patients remains largely unknown. Functional magnetic resonance imaging (fMRI) is a highly effective approach for evaluating and tracking neuropsychiatric diseases, such as bipolar disorder, schizophrenia, PD, and Alzheimer’s disease ^32–39^. Previous studies have identified a multitude of neuroimaging alterations related to the clinical characteristics of neuropsychiatric diseases through the analysis of functional and structural networks ^34,36,40^. Although there is little consensus in the fMRI signatures and correlates of anxiety in PD according to previous literature, recent researches suggest that anxiety is specifically associated with structural and functional alterations of both fear and limbic cortico-striato-thalamocortical circuits ^41^. Particularly, it has been shown that anxiety is associated with the structural and functional changes of amygdala ^42,43^. For example, lower left amygdala volume has been shown to contribute to the concomitant development of anxiety symptoms ^44^. Additionally, the functional connectivity between amygdala and hippocampus is negatively correlated with anxiety symptoms ^43^. However, it seems that the development of anxiety in PD is associated with more widespread abnormal activities of multiple brain regions, such as frontal cortex, basal ganglia, cerebellum, and limbic system ^45^. Therefore, the occurrence of anxiety in PD may be associated with multiple brain networks. Actually, PD patients with anxiety exhibit reduced functional connectivity within sensorimotor network (SMN) and default-mode network (DMN) and elevated functional connectivity within the executive-control network (ECN) compared to patients without anxiety ^46^. In addition, anxiety was also linked with abnormal cerebellum functional network, specifically involved in cerebellum, caudate, and anterior cingulate cortex ^47^. Structural network has been also reported to be associated with anxiety in PD. For example, it has been shown that severity of anxiety was negatively correlated with structural covariance between the left striatal sub-regions and the contralateral caudate nucleus ^48^. Moreover, PD patients with anxiety seem to have lower white matter integrity within the striato-orbitofrontal, cingulate-limbic, and caudate-thalamic tracts, which are all key components of brain fear circuit^49^. These studies suggest that fMRI is an excellent tool that can assist to identify the underlying neural mechanisms associated with anxiety in PD.

Recently, we revealed that brain topological metrics were significantly associated with clinical symptoms of PD patients and mediated the effects of age, sex, and disease-associated risk variant on clinical manifestations in PD ^50,51^. However, whether brain network topology is correlated with anxiety and mediates the effects of anxiety on clinical features of PD patients remain poorly understood. In this study, we hypothesize that topological changes of brain networks may be associated with anxiety and mediate the associations between anxiety and motor and non-motor symptoms. Specifically, the objectives of current study include: (i) to examine whether anxiety is associated with clinical symptoms of PD patients; (ii) to assess how anxiety shapes brain structural network metrics using graphical analysis; (iii) to explore whether brain network metrics mediate the effects of anxiety on motor and non-motor symptoms of PD patients using mediation analysis.

## Materials and Methods

### Participants

The data used in this study were obtained from the Parkinson’s Progression Markers Initiative (PPMI) database ^52,53^, accessible at www.ppmi-info.org. This PPMI dataset comprised more than 400 patients diagnosed with early PD and 200 participants serving as controls. They underwent standardized evaluations including imaging, biochemical, clinical, and behavioral assessments in order to advance the research on biomarkers linked to the progression of PD. The Institutional Review boards of each participating center approved all procedures, and all participants signed informed consents before participating the study. The criteria for qualifying PD patients in this study were as follows: (i) They were over 30 years old; (ii) They were diagnosed with PD according to the MDS Clinical Diagnostic Criteria for PD ^54^; (iii) They received 3D T1-weighted MPRAGE imaging and diffusion tensor imaging (DTI) within the same period. The participants were excluded if they exhibited noticeable abnormalities in T1-weighted or T2-weighted MRI scans, or carried genetic mutations associated with familial PD, or were part of the genetic PPMI cohort and prodromal cohort. According to above criteria, a total of 146 PD patients were included in the analysis. They received motor assessments including Hoehn and Yahr stages, Tremor scores, Total Rigidity scores, and MDS Unified Parkinson’s Disease Rating Scale (MDS-UPDRS). The non-motor assessments consisted of Epworth Sleepiness Scale (ESS), REM Sleep Behavior Disorder Screening Questionnaire (RBDSQ), 15-item Geriatric Depression Scale (GDS), Scale for Outcomes in Parkinson’s Disease-Autonomic (SCOPA-AUT), State-Trait Anxiety Inventory (STAI), Semantic Fluency Test (SFT), Benton Judgment of Line Orientation test (BJLOT), Letter Number Sequencing test (LNS), Symbol Digit Modalities Test (SDMT), and Montreal Cognitive Assessment (MoCA). In order to evaluate the effects of anxiety on clinical features and brain networks, PD participants (n = 146) were classified into lower quartile group (Q1, STAI rank: 0%∼25%, STAI score range: 40∼51), interquartile group (Q2-3, STAI rank: 26%∼75%, STAI score range: 51∼78), and upper quartile group (Q4, STAI rank: 76%∼100%, STAI score range: 78∼121) based on their STAI score quartiles. The clinical characteristics of PD patients in 3 quartile groups were shown in Table S1 and Figure 1.

**Figure 1.**
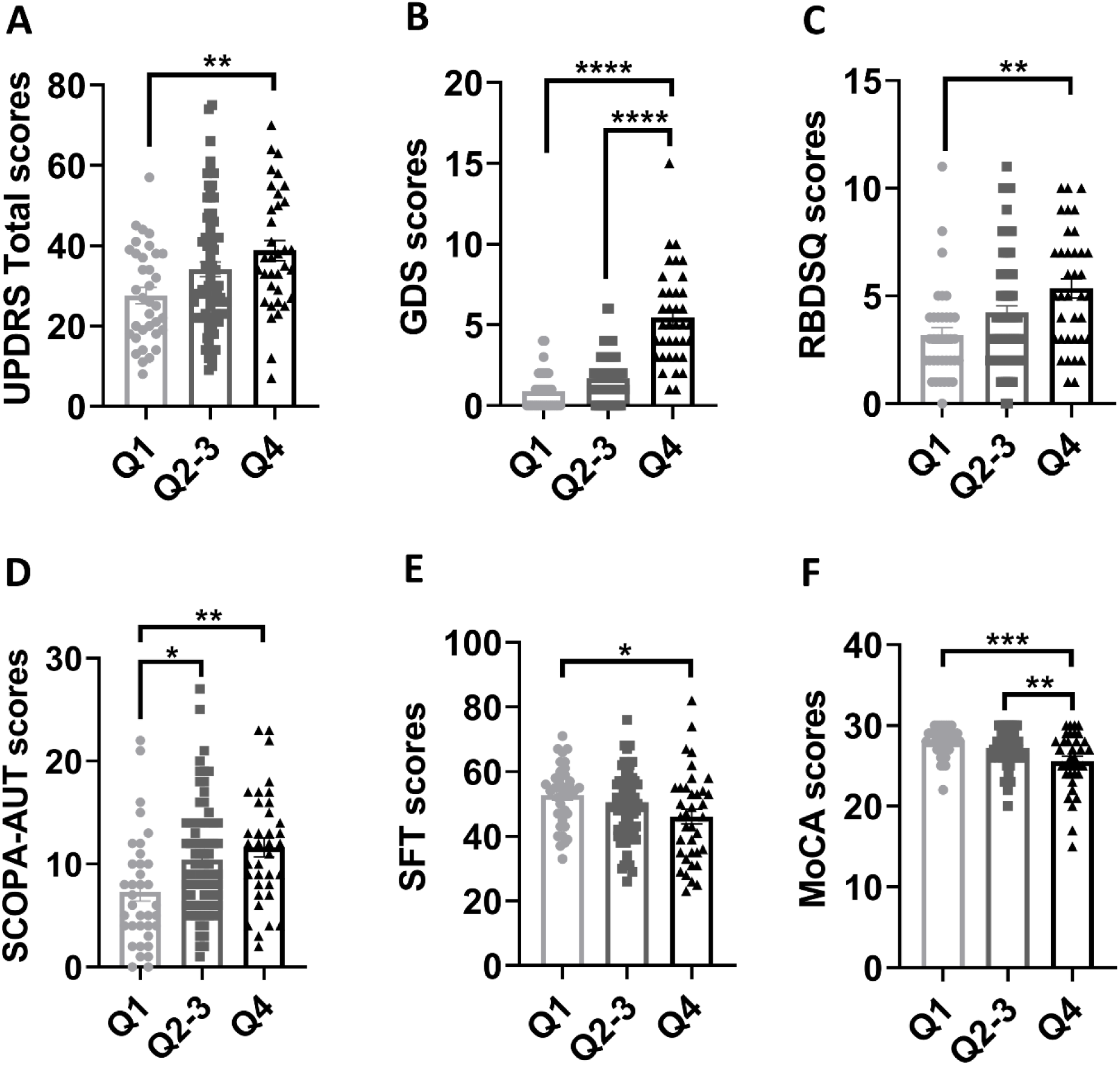
Group differences in clinical assessments. (A-F) Group differences in total UPDRS scores (A), GDS scores (B), RBDSQ scores (C), SCOPA-AUT scores (D), SFT scores (E), and MoCA scores (F). One-way ANOVA followed by Tukey’s post hoc test (Q1 group *vs* Q2-3 group *vs* Q4 group) were conducted to compare clinical variables. *p* < 0.05 was considered statistically significant. \**p* < 0.05, \*\**p* < 0.01, \*\*\**p* < 0.001, \*\*\*\**p* < 0.0001. Abbreviations: UPDRS, Unified Parkinson’s Disease Rating Scale; RBDSQ, REM Sleep Behavior Disorder Screening Questionnaire; SCOPA-AUT, Scale for Outcomes in Parkinson’s Disease-Autonomic; GDS, 15-item Geriatric Depression Scale; SFT, Semantic Fluency Test; MoCA, Montreal Cognitive Assessment.

### Image acquisition

The structural imaging data were acquired with Siemens 3T Trio or Verio scanners from Siemens Medical Solutions in Malvern, PA. The 3D T1-weighted MRI images were obtained using a magnetization-prepared rapid acquisition gradient echo sequence: Repetition time (TR) = 2300 ms, Echo time (TE) = 2.98 ms, Voxel size = 1 mm^3^, Slice thickness = 1.2 mm, twofold acceleration, and sagittal-oblique angulation. The DTI was scanned with the following protocol settings: TR = 8,400-8,800 ms, TE = 88 ms, Voxel size = 2 mm^3^, Slice thickness = 2 mm, 64 diffusion-sensitive gradient directions at b = 1,000 s/mm^2^ and one diffusion-unweighted (b0) image.

### Imaging preprocessing

The FMRIB Software Library toolkit (FSL, https://fsl.fmrib.ox.ac.uk/fsl/fslwiki) was used to preprocess DTI images of 146 PD patients. Briefly, the DTI images initially underwent corrections for head motions, eddy-current distortions, and susceptibility artifacts. Then, regular DTI metrics, such as fractional anisotropy (FA), mean diffusivity (MD), axial diffusivity, and radial diffusivity, were computed. Afterwards, the processed images were further reconstructed in the standard Montreal Neurological Institute (MNI) space for the generation of white matter networks.

### Network construction

A MATLAB toolkit, PANDA (http://www.nitrc.org/projects/panda/), was used to execute deterministic fiber tractography to construct structural networks. In short, Fiber Assignment by Continuous Tracking (FACT) algorithm was performed to generate white matter fibers connecting all 90 cortical and subcortical nodes in the Automated Anatomical Labeling (AAL) atlas. The FA threshold was set to 0.20, and the fiber angle threshold was set to 45 °. After white matter tractography, a 90 x 90 structural network based on fiber number (FN) was generated for individual participant.

### Graph-based network analysis

The topological properties of structural networks were calculated with GRETNA toolbox (https://www.nitrc.org/projects/gretna/) ^55^. A range of network sparsity thresholds (0.05 ∼ 0.50 with an interval of 0.05) was applied to compute both the global and nodal network metrics. The global network properties consisted of global efficiency, local efficiency, and small-worldness properties. The nodal network metrics included nodal betweenness centrality (BC), nodal degree centrality (DC), nodal clustering coefficient (Cp), nodal efficiency, nodal local efficiency, and nodal shortest path length. The detailed definitions for all network measurements have been documented by previous studies ^56–58^.

### Statistical analysis

#### Comparison of clinical variables

One-way ANOVA followed by Tukey’s post hoc test (Q1 group *vs* Q2-3 group *vs* Q4 group) was performed to evaluate the group differences of continuous variables. The categorical variables were analyzed using χ^2^ test. *p* < 0.05 was considered statistically significant.

#### Comparison of global network strength

The comparisons of global network strength of brain networks were performed using Network-Based Statistic (NBS, https://www.nitrc.org/projects/nbs/) toolbox. *p* < 0.05 after false discovery rate (FDR) correction was considered statistically significant. During NBS analysis, age, sex, years of education, and disease duration were enrolled as covariates.

#### Comparison of network metrics

The comparisons of global and nodal network metrics were conducted using two-way ANOVA test followed by FDR corrections. *p* < 0.05 after FDR correction was considered statistically significant.

#### Association analysis

The associations between STAI scores and clinical variables or between clinical variables (i.e., STAI scores) and network metrics were performed with both Pearson correlation method and multivariate regression analysis. During multivariate regression analysis, age, sex, disease duration, and education level, were included as covariates. *p* < 0.05 was considered statistically significant for associations between STAI scores and clinical variables. *p* < 0.05 after FDR correction was considered statistically significant for associations between clinical variables and graphical network metrics.

### Mediation analysis

Mediation analysis was performed using IBM SPSS Statistics Version 26. STAI scores were entered as independent variable during mediation analysis. The dependent variables were the scores of clinical assessments (UPDRS-III, ESS, SFT and MoCA). The mediators were graphical network metrics. The indirect effects of graphical metrics on the associations between STAI scores and clinical assessments were modelled. The covariates included age, sex, disease duration, and years of education. *p* < 0.05 was considered statistically significant for mediation analysis.

## Results

### Group differences in clinical variables

The Age, sex, years of education, and disease duration were not statistically different among 3 quartiles (Table 1). Compared to Q1 group (n = 36), Q2-3 (n = 73) and Q4 group (n = 37) exhibited higher STAI scores (both *p* < 0.0001). Compared to Q1 group, Q4 group showed higher total UPDRS scores (*p* < 0.01; Figure 1A), GDS scores (*p* < 0.0001; Figure 1B), RBDSQ scores (*p* < 0.01; Figure 1C), SCOPA-AUT scores (*p* < 0.01; Figure 1D), and lower SFT (*p* < 0.05; Figure 1E) and MoCA scores (*p* < 0.001; Figure 1F).

**TABLE 1.** The demographic and clinical data for each quartile group The data were shown as the mean ± standard deviation (SD).

| Clinical variable | Q1 group<br>(n = 36) | Q2-3 group<br>(n = 73) | Q4 group<br>(n = 37) | Q1 vs Q2-3 | Q1 vs Q4 | Q2-3 vs Q4 |
| --- | --- | --- | --- | --- | --- | --- |
| Age, years | 60.41 ± 8.76 | 61.84 ± 9.24 | ± | *p* = 0.7291 | *p* = 0.9769 | *p* = 0.8586 |
| Sex (Male/Female) | 22/14 | 50/23 | 21/16 | *p* = 0.4440 | *p* = 0.7054 | *p* = 0.2241 |
| Education, years | 15.22 ± 3.15 | 15.48 ± 2.78 | 14.89 ± 3.25 | *p* = 0.9069 | *p* = 0.8852 | *p* = 0.5964 |
| Disease duration, years | 2.08 ± 2.08 | 2.12 ± 2.50 | 1.83 ± 1.35 | *p* = 0.9961 | *p* = 0.8664 | *p* = 0.7775 |
| UPDRS-III scores | 19.40 ± 9.92 | 20.73 ± 9.38 | 22.57 ± 9.90 | *p* = 0.7820 | *p* = 0.3564 | *p* = 0.6218 |
| STAI scores | 44.97 ± 3.44 | 63.27 ± 7.47 | 93.03 ± 11.79 | ***p* < 0.0001** | ***p* < 0.0001** | ***p* < 0.0001** |

The data were shown as the mean ± standard deviation (SD). One-way ANOVA followed by Tukey’s post hoc test (Q1 group vs Q2-3 group vs Q4 group) were utilized to compare continuous variables. $\chi^2$ test was used to compare categorical variable (Sex). *p* < 0.05 was considered statistically significant. The bold values indicate statistical significance. Abbreviations: STAI, State-Trait Anxiety Inventory.

### Associations between STAI scores and clinical variables

To evaluate whether the effects of anxiety severity on clinical assessments were independent of age, sex, disease duration, and years of education, we analyzed the associations between anxiety severity and clinical assessments using multivariate regression analysis. As shown in Table 2, STAI scores were positively associated with UPDRS-III scores, total UPDRS scores, ESS scores, GDS scores, RBDSQ scores, SCOPA-AUT scores, and negatively associated with scores of SFT, LNS, MoCA, Immediate Recall and Delayed Recall of HVLT-R (Table 2).

**TABLE 2.** Associations between STAI scores and clinical assessments.

| Clinical assessments |  |  |  |  |  |  |  |  |
| --- | --- | --- | --- | --- | --- | --- | --- | --- |
| Variables<br>Statistics | HY stage | Tremor | Rigidity | UPDRS-III | UPDRS-Total | ESS | GDS | RBDSQ |
| $\beta$ value | $\beta = \mathbf{0.007}$ | $\beta = -0.01$ | $\beta = 0.02$ | $\beta = \mathbf{0.10}$ | $\beta = \mathbf{0.28}$ | $\beta = \mathbf{0.06}$ | $\beta = \mathbf{0.09}$ | $\beta = \mathbf{0.05}$ |
| $p$-value | $p = \mathbf{0.0016}$ | $p = 0.3753$ | $p = 0.1826$ | $p = \mathbf{0.0181}$ | $p < \mathbf{0.0001}$ | $p = \mathbf{0.0007}$ | $p < \mathbf{0.0001}$ | $p < \mathbf{0.0001}$ |
| Variables<br>Statistics | SCOPA-AUT | BJLOT | SFT | SDMT | LNS | MoCA | HVLT-R<br>Immediate Recall | HVLT-R<br>Delayed Recall |
| $\beta$ value | $\beta = \mathbf{0.09}$ | $\beta = -0.01$ | $\beta = \mathbf{-0.10}$ | $\beta = \mathbf{-0.09}$ | $\beta = \mathbf{-0.02}$ | $\beta = \mathbf{-0.04}$ | $\beta = \mathbf{-0.04}$ | $\beta = \mathbf{-0.03}$ |
| $p$-value | $p = \mathbf{0.0003}$ | $p = 0.1089$ | $p = \mathbf{0.0306}$ | $p = \mathbf{0.0391}$ | $p = \mathbf{0.0401}$ | $p < \mathbf{0.0001}$ | $p = \mathbf{0.0460}$ | $p = \mathbf{0.0154}$ |

The data were shown as the $\beta$ and $p$ -values derived from multivariate regression analysis with age, sex, years of education, and disease duration as covariates. The motor function examination was assessed in ON state. $p < 0.05$ was considered statistically significant. The bold values indicate statistical significance. Abbreviations: HY, Hoehn & Yahr; UPDRS, Unified Parkinson’s Disease Rating Scale; ESS, Epworth Sleepiness Scale; RBDSQ, REM Sleep Behavior Disorder Screening Questionnaire; GDS, Geriatric Depression Scale; SCOPA-AUT, Scale for Outcomes in Parkinson's Disease-Autonomic; SDMT, Symbol Digit Modalities Test; LNS, Letter Number Sequencing; SFT, Semantic Fluency Test Score; STAI, State-Trait Anxiety Inventory; BJLOT, Benton Judgement of Line Orientation; MoCA, Montreal Cognitive Assessment; HVLT-R, Hopkins Verbal Learning Test-Revised.

### Group differences in structural networks

To examine whether anxiety shaped structural network metrics, we compared the group differences of graphical network metrics among 3 quartile groups. For global network metrics in structural network, no significant difference was observed among 3 quartile groups. For nodal BC, Q4 group showed higher nodal BC in right superior temporal gyrus, left middle temporal gyrus, and right middle temporal gyrus (FDR-corrected *p* < 0.05; Figure 2A). Compared to Q1 group, Q4 group exhibited lower nodal BC (FDR-corrected *p* < 0.05 in left calcarine, left superior parietal gyrus, left thalamus, left middle temporal pole; Figure 2B), nodal Cp (FDR-corrected *p* < 0.05 in left middle orbitofrontal gyrus and left inferior occipital gyrus; Figure 2C), nodal efficiency (FDR-corrected *p* < 0.05 in left calcarine and left superior parietal gyrus; Figure 2D), nodal local efficiency (FDR-corrected *p* < 0.05 in left middle orbitofrontal gyrus, right amygdala, and left inferior occipital gyrus; Figure 2E), and nodal shortest path length (FDR-corrected *p* < 0.05 in right rectus; Figure 2F) in structural network.

**Figure 2.**
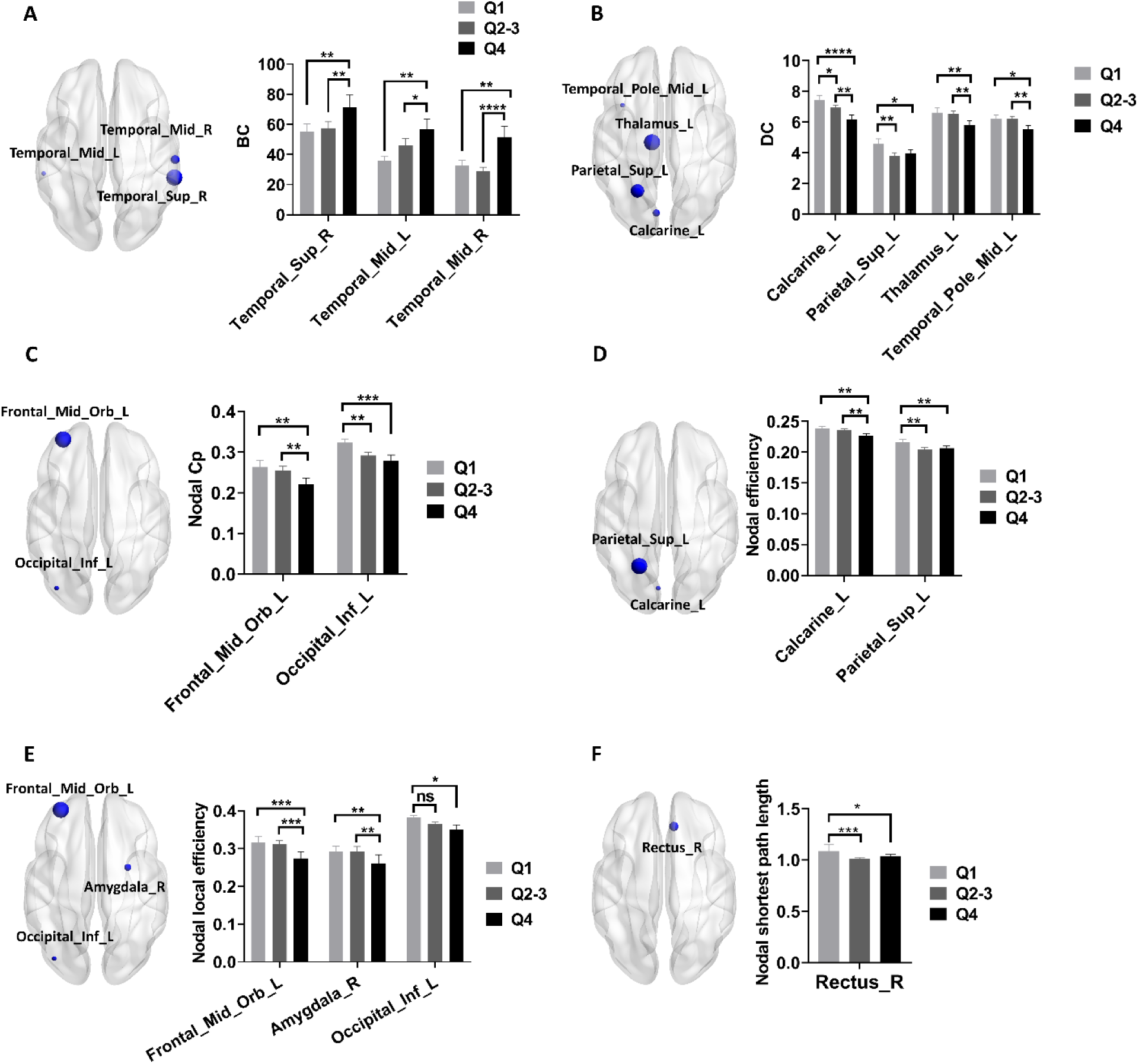
Group differences of nodal network metrics in structural network. (A-D) Group differences of nodal BC (A), nodal DC (B), nodal Cp (C), nodal efficiency (D), nodal local efficiency (E), and nodal shortest path length (F) in structural network among 3 quartile groups. Two-way ANOVA test with FDR correction was performed. *p* < 0.05 after FDR correction was considered to have statistical significance. \**p* < 0.05, ** *p* < 0.01, \*\*\**p* < 0.001, \*\*\*\**p* < 0.0001. Abbreviations: BC, Betweenness centrality; DC, degree centrality; Cp, Clustering coefficient.

### Group differences in global network strength

With NBS analysis, we found no significant differences in global network strength of structural network among 3 quartile groups (FDR-corrected *p* > 0.05, NBS method).

### Associations between STAI scores and graphical network metrics

As shown in Table 3, STAI scores were associated with BC in right middle temporal gyrus, DC in left calcarine, nodal Cp in left inferior occipital gyrus, nodal efficiency in left calcarine, and nodal local efficiency in left inferior occipital gyrus (FDR-corrected *p* < 0.05; Table 3).

**TABLE 3.**
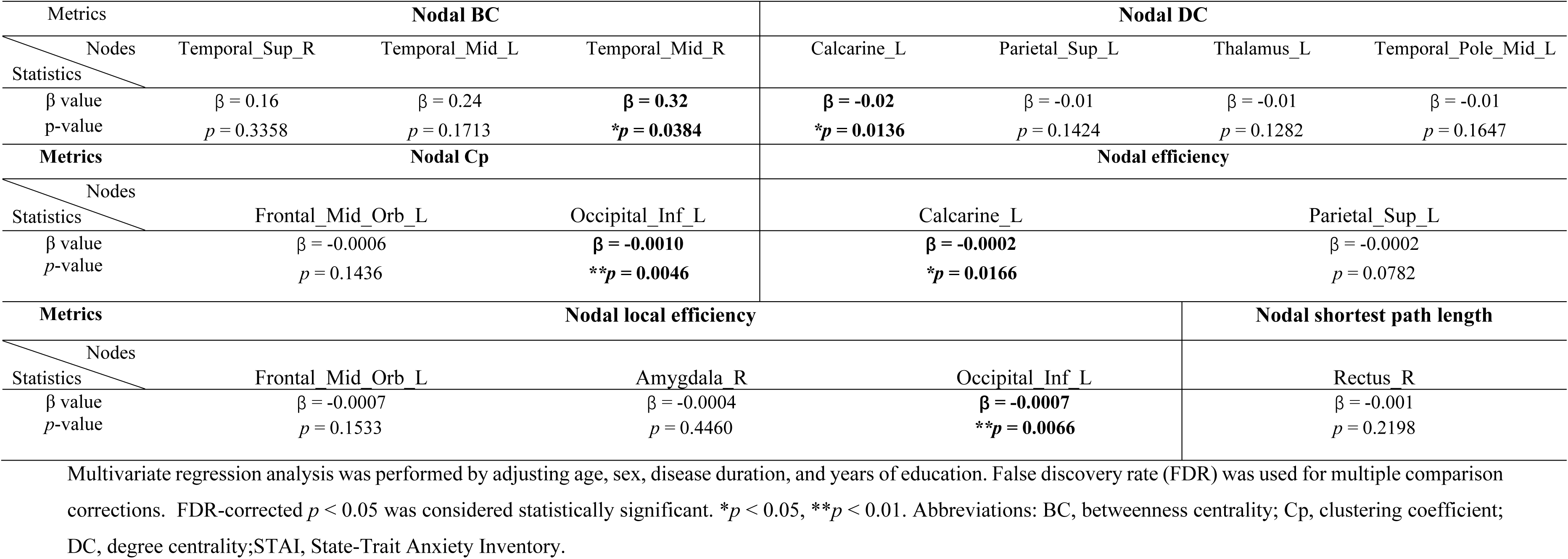
The associations between STAI scores and structural network metrics.

### Mediation analysis for motor symptoms

We revealed STAI scores were positively associated with UPDRS-III scores (β = 0.10, *p* = 0.0181; Table 1) and STAI scores were associated with multiple structural network metrics (Table 3), therefore, we examined whether brain structural network metrics mediated the effects of anxiety on UPDRS-III scores. As shown in Figure 3, we initially showed that DC of left calcarine, nodal Cp of left inferior occipital gyrus, nodal efficiency of left calcarine, and nodal local efficiency of left inferior occipital gyrus were negatively associated with UPDRS-III scores (FDR-corrected *p* < 0.05; Figure 3A-D). Then, with mediation analysis, we further demonstrated that both DC and nodal efficiency of left calcarine mediated the positive association between STAI scores and UPDRS-III scores (Figure 3E-F). The mediation analysis of nodal Cp and nodal local efficiency of left inferior occipital gyrus showed no statistical significance.

**Figure 3.**
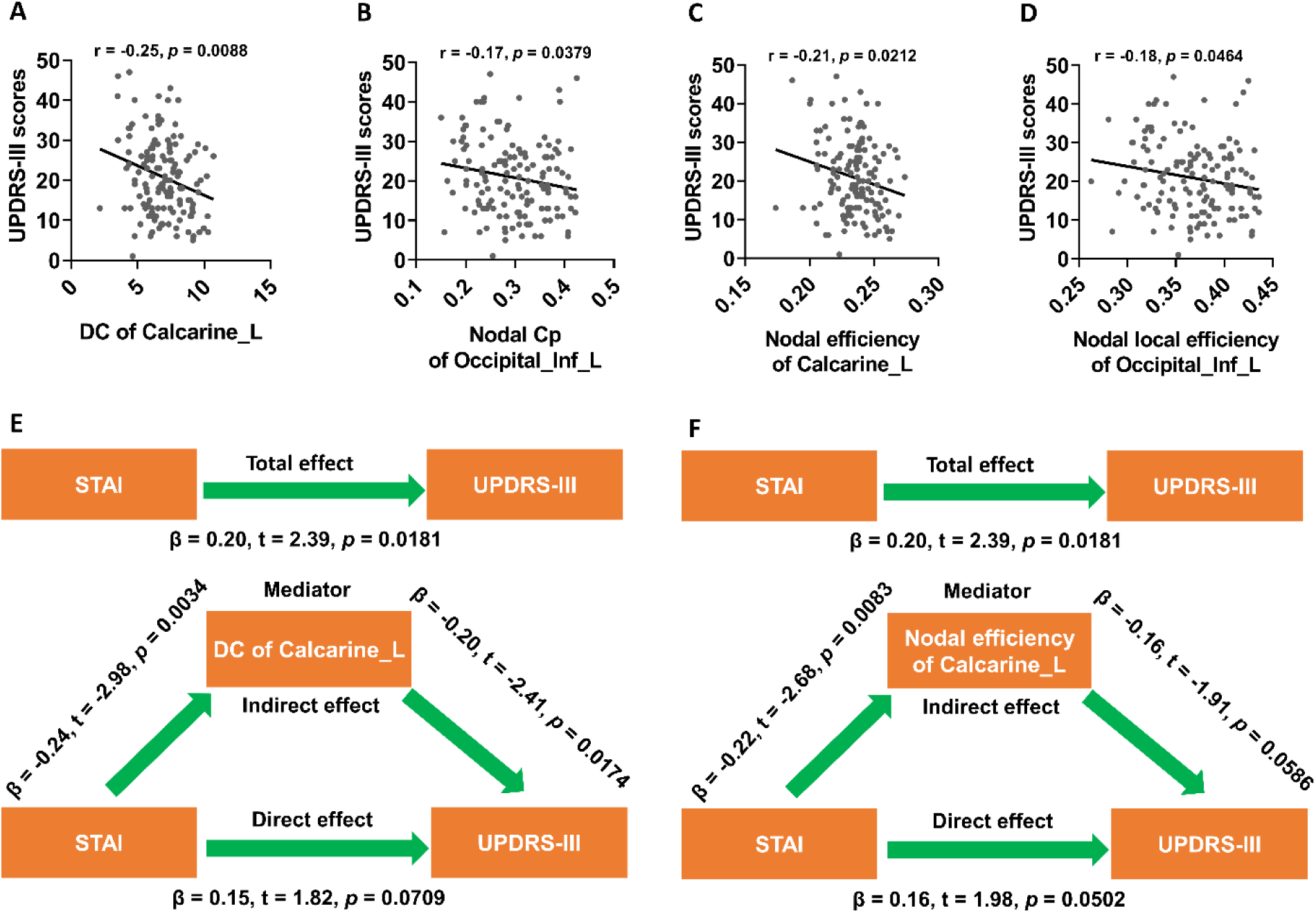
Nodal DC and efficiency of left calcarine mediated the association between anxiety and UPDRS-III scores. (A-D) DC of left calcarine, nodal Cp of left inferior occipital gyrus, nodal efficiency of left calcarine, and nodal local efficiency of left inferior occipital gyrus were negatively associated with UPDRS-III scores (FDR-corrected *p* < 0.05). (E-F) Nodal DC and efficiency of left calcarine mediated the positive associations between anxiety and UPDRS-III scores. The associations between UPDRS-III scores and network metrics were performed with Pearson correlation method (FDR-corrected *p*-values were shown). During the mediation analysis, age, sex, disease duration, and years of education were included as covariates. *p* < 0.05 was considered statistically significant. Abbreviations: DC, Degree centrality; Cp, Clustering coefficient; UPDRS, Unified Parkinson’s Disease Rating Scale; STAI, State-Trait Anxiety Inventory.

### Mediation analysis for non-motor symptoms

We found STAI scores were positively associated with ESS scores (β = 0.06, *p* = 0.0007; Table 2) and DC of left calcarine was negatively associated with ESS scores (Figure 4A). Thus, we assessed whether DC of left calcarine mediated the association between STAI scores and ESS scores. Indeed, we found DC of left calcarine mediated the positive association between STAI scores and ESS scores (Figure 4B). Consistently, we revealed nodal efficiency of left calcarine was negatively associated with ESS scores (Figure 4C) and mediated the positive association between STAI scores and ESS scores (Figure 4D).

**Figure 4.**
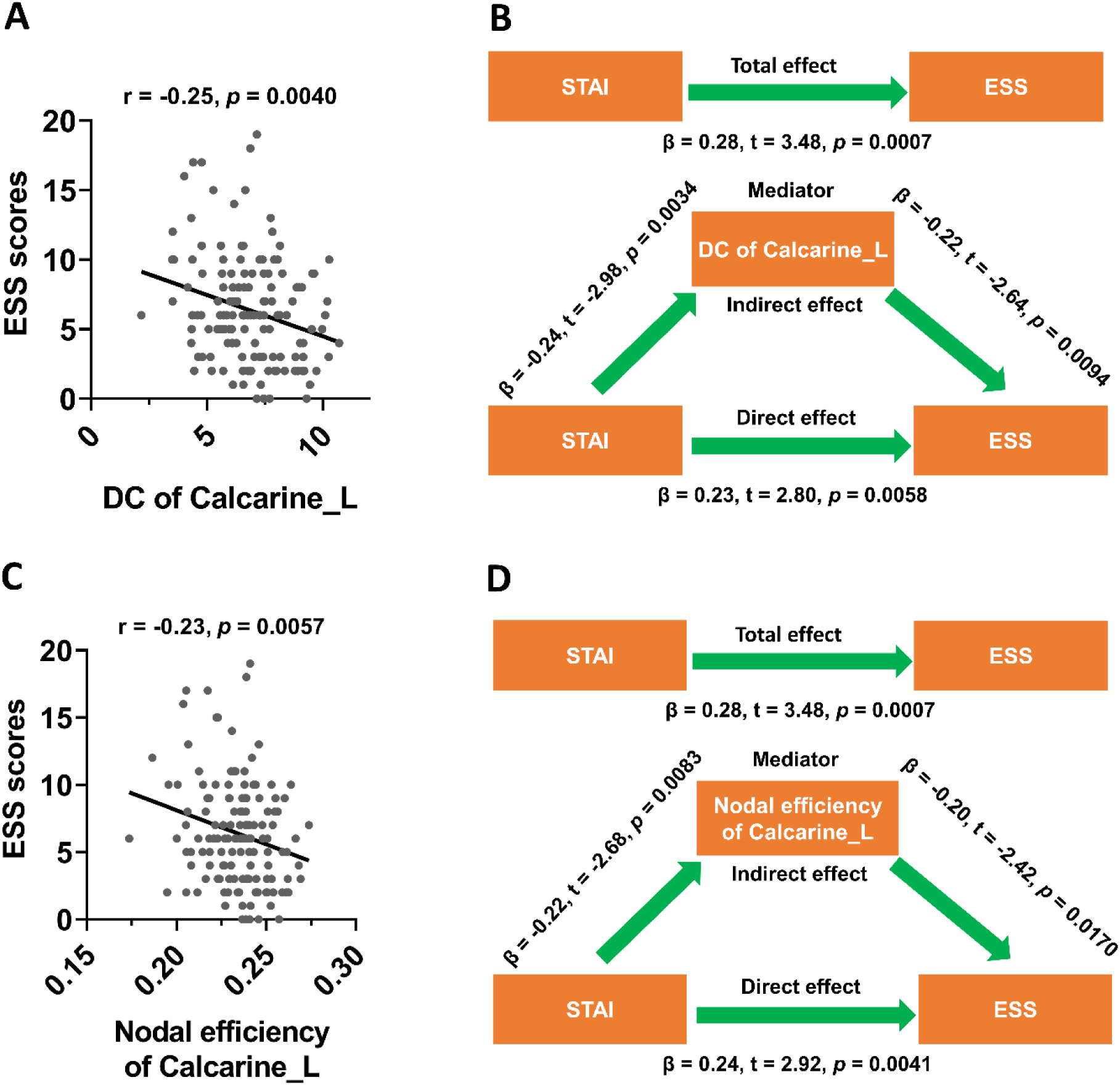
Nodal DC and efficiency of left calcarine mediated the association between anxiety and ESS scores. (A) Nodal DC of left calcarine was negatively associated with ESS scores. (B) Nodal DC left calcarine mediated the positive associations between anxiety and ESS scores. (C) Nodal efficiency of left calcarine was negatively associated with ESS scores. (D) Nodal efficiency left calcarine mediated the positive associations between anxiety and ESS scores. The associations between ESS scores and network metrics were performed with Pearson correlation method (FDR-corrected *p*-values were shown). During the mediation analysis, age, sex, disease duration, and years of education were included as covariates. *p* < 0.05 was considered statistically significant. Abbreviations: DC, Degree centrality; ESS, Epworth Sleepiness Scale; STAI, State-Trait Anxiety Inventory.

In the Table 2, we found STAI scores were negatively associated with cognitive function of PD patients (Table 2), thus, we hypothesized that structural network metrics may mediate the associations between STAI scores and cognitive function of the patients. In the Figure 5, we found BC in right middle temporal gyrus was negatively associated with SFT scores (FDR-corrected *p* < 0.05; Figure 5A), and additionally BC in right middle temporal gyrus (FDR-corrected *p* < 0.05; Figure 5B), nodal Cp in left inferior occipital gyrus (FDR-corrected *p* < 0.05; Figure 5C), and nodal efficiency in left calcarine (FDR-corrected *p* < 0.05; Figure 5D) were negatively associated with MoCA scores. During mediation analysis, we demonstrated that BC in right middle temporal gyrus mediated the negative association between STAI scores and SFT scores (Figure 5E), and BC in right middle temporal gyrus, nodal Cp in left inferior occipital gyrus, and nodal efficiency in left calcarine partially mediated negative association between STAI scores and MoCA scores (Figure 5F-H).

**Figure 5.**
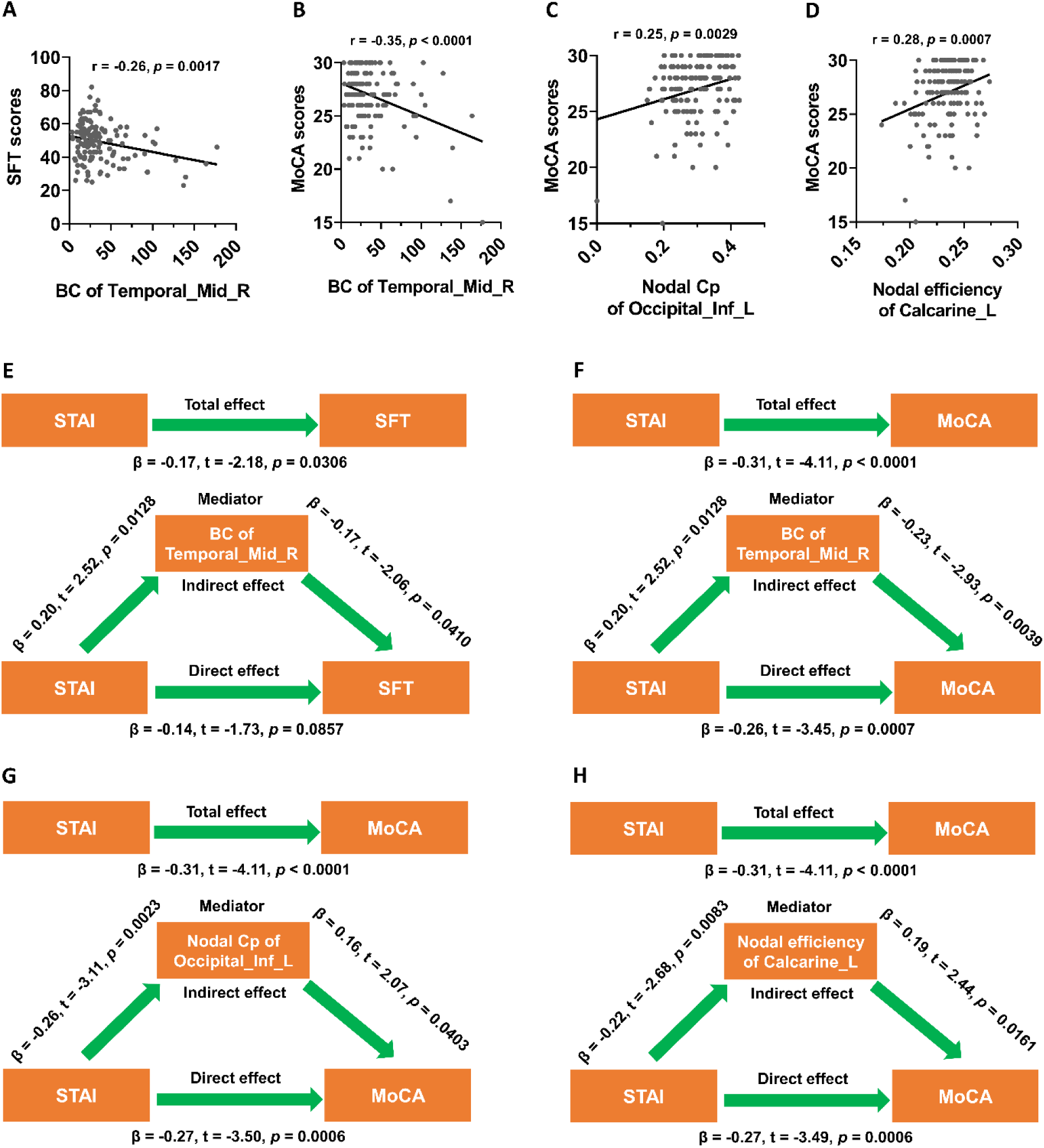
Structural network metrics mediated the associations between anxiety and cognitive function. (A-B) Nodal BC of right middle temporal gyrus was negatively associated with SFT and MoCA scores (FDR-corrected *p* < 0.05). (C-D) Nodal Cp of left inferior occipital gyrus and nodal efficiency of left calcarine were positively associated with MoCA scores (FDR-corrected *p* < 0.05). (E-F) Nodal BC of right middle temporal gyrus mediated the negative associations between anxiety and SFT and MoCA scores. (G-H) Nodal Cp of left inferior occipital gyrus and nodal efficiency left calcarine mediated the negative associations between anxiety and MoCA scores. The associations between SFT or MoCA scores and network metrics were performed with Pearson correlation method (FDR-corrected *p*-values were shown). During the mediation analysis, age, sex, disease duration, and years of education were included as covariates. *p* < 0.05 was considered statistically significant. Abbreviations: BC, Betweenness centrality; Cp, Clustering coefficient; SFT, Semantic Fluency Test; MoCA, Montreal Cognitive Assessment; STAI, State-Trait Anxiety Inventory.

## Discussion

In this study, we replicated previous studies that anxiety was associated with both motor and non-motor features in PD patients. In addition, with graphical analysis, we revealed anxiety significantly modified the network topology of brain structural networks. Furthermore, we observed significant associations between differential graphical network metrics and anxiety-associated clinical manifestations. Finally, we demonstrated that graphical network metrics mediated the associations between anxiety and motor and non-motor symptoms.

### 1. The effects of anxiety on clinical phenotypes of PD patients

Recently, there has been a growing focus on the impact of anxiety on clinical severity and quality of life in PD patients. In our study, we found anxiety was positively associated with HY stages, indicating that anxiety was associated with worse disease severity, which was consistent with a previous finding that higher HY stages was associated with increased anxiety ^59^. In agreement with this finding, we also revealed that anxiety was positively associated with higher UPDRS-III scores and total UPDRS scores, which is independent of age, sex, disease duration, and years of education. In consistent with above results, a previous study has shown that Beck Anxiety Inventory (BAI) total scores were significantly correlated with poorer UPDRS motor score ^60^. In addition to motor symptoms, previous studies also showed that anxiety was associated with non-motor symptos, such as EDS ^11^, depression ^5,28,29^, RBD ^12,13^, and autonomic dysfunction ^12,28^, which were consistent with our findings shown in Table 2. According to previous literature, anxiety was associated with cognitive decline in PD across multiple cognitive domains, including attention/working memory, executive function, and language ^21^. In addition, higher level of anxiety was associated with impairment of immediate verbal memory ^24^. Moreover, social anxiety was also associated with negative social cognitions and safety-seeking behaviors ^61^. Importantly, anxiety level was found to be associated with the development of mild cognitive impairment in PD patients ^20^. Consistently, we found anxiety was negatively associated with scores of multiple cognitive scales, especially global cognition and verbal memory. Taken together, our findings suggested that anxiety was significantly associated with the severity of motor and non-motor manifestations, which would dramatically decline the quality of life in PD patients ^3,5^.

### 2. The effects of anxiety on structural network

Although anxiety was associated with multiple clinical features, the underlying neural mechanisms were still unknown. To decipher the neural mediators of anxiety that contribute to its effects on clinical symptoms, we assessed how anxiety modulated brain structural networks. Though NBS analysis showed that global network strengths in brain structural networks were not statistically different among 3 quartiles, we found anxiety significantly shaped the topological metrics of brain networks. For nodal BC, we found higher nodal BC in higher level of anxiety group (Q4 group) compared to lower level of anxiety group (Q1 group). Nodal BC measures the number of times a node passing through the shortest path between other nodes, which displays which nodes are “bridges” or “connectors” between nodes within a network ^56,62^. Therefore, the increase of nodal BC in patients with higher level of anxiety indicates that bilateral temporal gyri become key “bridges” in structural network of these patients, which may be associated the changes of structural connectivity. Using multivariate regression analysis, we further revealed that nodal BC of right middle temporal gyrus was independent of anxiety, indicating anxiety specifically modified the local topology of right middle temporal gyrus. We found nodal efficiency and nodal local efficiency were significantly lower in higher level of anxiety group, indicating that higher anxiety impaired the local information transfer of structural network, which may be due to the reductions of nodal DC and nodal Cp in higher level of anxiety group. Thus, these results suggested that higher level of anxiety induced dramatical decline of information transfer efficiency in structural network. With multivariate regression analysis, we further showed that STAI scores were independently associated with some of these network metrics, indicating that the effects of anxiety on network topology is independent of other confounding variables, including age and sex.

Interestingly, we found the nodal network metrics of left calcarine and left inferior occipital gyrus were independently associated with anxiety, which indicated that anxiety specifically shaped the local network topology of left occipital areas.

### 3. The associations between anxiety and motor symptoms

In our study, anxiety was significantly associated with HY stage, UPDRS-III scores, and total UPDRS scores, which suggested that high level of anxiety was associated with worse motor symptoms. Therefore, our findings were consistent with the results reported by previous studies ^59,60^. In the Figure 2, we showed both lower nodal DC and lower nodal efficiency of left calcarine in high level of anxiety group compared to low level of anxiety group, which was independent of age, sex, and disease duration. In a recent study, we revealed that nodal DC of left calcarine was negatively associated with UPDRS-III scores and mediated the effects of autonomic dysfunction on UPDRS-III scores ^63^. Therefore, we examined whether nodal DC and nodal efficiency of left calcarine were also associated with UPDRS-III scores in this study. Indeed, nodal DC and nodal efficiency of left calcarine were negatively associated with UPDRS-III scores. With mediation analysis, we further demonstrated that nodal DC and nodal efficiency of left calcarine mediated the positive association between anxiety and UPDRS-III scores. These results were supported by a recent study showing that DC at the left calcarine was negatively associated with scores of UPDRS-III in PD patients ^64^. As a key node in visual network, the function and structure of left calcarine have been found to be impaired in PD. For example, PD patients showed decreased regional homogeneity in left calcarine compared to healthy control ^65^. In addition, it has also been shown that lower gray matter volume in the left calcarine correlated with poor attention and executive function, which exacerbated motor function in PD patients ^66^. Therefore, it is possible that the dysfunction of left calcarine may contribute to impaired cognitive processing in PD, which resulted in aberrant motor control and motor deterioration. Actually, we showed that nodal efficiency of left calcarine mediated the effects of anxiety on MoCA scores. In agreement with this hypothesis, previous researches have demonstrated left calcarine was associated with cognition decline in multiple diseases. For instance, in AD, reduced gray matter volume of left calcarine was correlated with the impairment of visuo-spatial cognition and episodic memory in *APOE* ε4 carriers ^67^. In addition, it has been shown that functional connectivity of left calcarine was negatively correlated with MoCA scores in Tinnitus patients with cognitive impairment ^68^. Furthermore, in type 2 diabetes mellitus, patients with cognitive impairment exhibited lower functional connectivity in the left calcarine, which was associated with lower episodic memory performance in these patients ^69^. Based on these findings, we concluded that left calcarine was a key node involved in cognitive function and mediated the effects of anxiety on motor and cognitive impairment of PD patients.

### 4. The associations between anxiety and EDS

In this study, we found anxiety was positively associated with ESS scores, which was consistent with the results reported by a previous study ^11^. How anxiety contributed to EDS is still unknown. One potential mechanism is that anxiety induced poor sleep quality at night in PD patients ^8^, which led to hypersomnia at daytime. In our study, we showed both nodal DC and nodal efficiency of left calcarine were negatively associated with ESS scores and mediated the positive association between anxiety and ESS scores. These results indicated that local topology of left calcarine contributed to EDS in PD patients, which was supported by our recent study ^70^. In fact, a series of studies have revealed the structure and function of left calcarine were significantly changed in patients with sleep disturbances. For example, the functional connectivity between left calcarine and right superior occipital gyrus was much lower in PD patients with RBD compared to patients without RBD ^71^. Additionally, the functional connectivity between left calcarine and locus coeruleus was significantly enhanced in chronic insomnia disorder ^72^. Moreover, it has been shown that gray matter volume of left calcarine was significantly reduced in poor sleepers of patients with cerebral small vessel disease (CSVD) compared to good sleepers of CSVD patients ^73^. Furthermore, gray matter volume in left calcarine cortex was associated with sleep quality in CSVD patients ^73^. Taken together, we concluded that changes in local topology of left calcarine was associated with EDS in PD patients.

### 5. The associations between anxiety and cognitive impairment

We observed anxiety was negatively associated with scores of multiple cognitive scales, especially global cognition and verbal memory, which have been demonstrated by several studies ^20,21,24,61^. We also revealed structural network metrics were significantly associated with cognitive function and mediated the effects of anxiety on cognitive function of PD patients. Therefore, here we provided network level of mechanisms to explain the negative association between anxiety and cognitive function. We showed BC of right middle temporal gyrus was significantly associated with SFT and MoCA scores, which was consistent with our recent findings showing that BC of right middle temporal gyrus mediated the negative association between age and SDMT scores ^74^. These results indicated that the structure and function of right middle temporal gyrus may be involved in cognitive impairment of PD patients. Indeed, lower gray matter volume in temporal lobe has been observed in PD patients with cognitive impairment compared to cognitively normal PD patients ^75^. In addition, Chiang *et al*. (2018) revealed that disruption of mesial temporal gyrus functional connectivity contributed to cognitive decline in PD ^76^. We revealed nodal network metrics of left inferior occipital gyrus and left calcarine were also associated with cognitive impairment of PD patients and mediated the effects of anxiety on cognitive function of PD patients. These results suggested that anxiety negatively impacted global cognitive function of PD patients by regulating the topology of multiple cognition-related regions located in temporal and occipital regions ^77–79^.

### 6. The molecular mechanisms underlying anxiety in PD

Although anxiety was an essential non-motor symptom in PD, its molecular basis remained largely elusive. According to previous studies, the disruptions of serotonergic, GABAergic, and adrenergic neural systems were all essential for the development of anxiety in PD ^6^. For example, 5-HT_1B_ receptor in the basolateral amygdala has been found to be involved in the occurrence of PD-related anxiety ^80^. ^123^I-N-ω-fluoropropyl-2β-carbomethoxy-3β-(4-iodophenyl) nortropane (^123^I-FP-CIT) is a radiotracer specifically binding serotonin transporter in extrastriatal subcortical regions ^81^. It has been demonstrated that higher level of anxiety in PD patients was associated with lower thalamic ^123^I-FP-CIT binding, implying an involvement of serotonergic degeneration in anxiety in PD ^81^. Dopamine neurotransmission also played an essential role in the occurrence of anxiety in PD patients. It was shown that impaired dopaminergic transmission in the globus pallidus increases anxiety-like behavior without altering motor activity ^82^. In addition, 1-methyl-4-phenyl-1,2,3,6-tetrahydropyridine (MPTP)-induced dopamine depletion in basolateral amygdala suppressed GABA_A_ receptors expression and long-term depression induction, which led to anxiety-like behaviors ^83^. As a key pathological signature in PD, α-synuclein also played an important role in the pathogenesis of anxiety in PD. It has been demonstrated that pathological α-synuclein propagation from the olfactory bulb to limbic system as well as its related regions led to the development of anxiety-like behavior in PD ^84^. Taken together, multiple molecular mechanisms were involved in the occurrence of anxiety in PD.

### 7. Strengths and limitations of this study

In this study, we revealed that high level of anxiety was associated with worse motor and non-motor symptoms, which emphasized the essential role of anxiety in clinical management of PD patients. We showed anxiety significantly shaped the nodal network metrics of structural network, which indicated that anxiety had important impacts on the structural network topology in PD patients. We demonstrated nodal network metrics of structural network mediated the associations between anxiety and motor and non-motor symptoms, implying that topological metrics in structural network were network mediators of anxiety-dependent motor and non-motor symptoms. Therefore, our findings provided network level mechanisms to interpret the key role of anxiety in the regulation of motor and non-motor features, which promoted the understanding of neural mechanisms underlying anxiety-dependent clinical phenotypes in PD. The limitation of this study was its cross-sectional design, future studies were required to examine whether nodal network topology contributed to the association anxiety and clinical manifestations in longitudinal cohort studies.

## Conclusions

Higher anxiety level is associated with worse clinical manifestations and aberrations of structural network topology. Differential structural network metrics mediated the associations between anxiety and motor and non-motor symptoms of PD patients.

## Author contributions

Zhichun Chen, Conceptualization, Formal analysis, Visualization, Methodology, Writing, review and editing; Guanglu Li, Data curation, Formal analysis, Visualization; Liche Zhou, Data curation, Formal analysis, Investigation; Lina Zhang, Formal analysis, Investigation, Methodology; Li Yang, Supervision, Writing, review and editing; Jun Liu, Conceptualization, Supervision, Funding acquisition, Writing, review, and editing.

## Acknowledgments

Data used in the preparation of this article were obtained from the Parkinson’s Progression Markers Initiative (PPMI) database (www.ppmiinfo.org/data). We thank the share of PPMI data by all the PPMI study investigators. PPMI – a public-private partnership – is funded by the Michael J. Fox Foundation for Parkinson’s Research and funding partners, which can be found at www.ppmiinfo.org/fundingpartners.

## Funding information

This work was supported by grants from National Natural Science Foundation of China (Grant No. 81873778, 82071415) and National Research Center for Translational Medicine at Shanghai, Ruijin Hospital, Shanghai Jiao Tong University School of Medicine (Grant No. NRCTM(SH)-2021-03).

## Conflict of Interest

The authors have no conflict of interest to report.

## Data availability

All the raw data used in the preparation of this Article were downloaded from PPMI database (www.ppmi-info.org/data).All data produced in the present study are available upon reasonable request to the authors.

## Supporting Information

Additional supporting information may be found online in the Supporting Information section at the end of the article.

## Notes

### Competing Interest Statement

The authors have declared no competing interest.

### Author Declarations

The Institutional Review boards of each participating center approved all procedures, and all participants signed informed consents before participating the study.

